# Young Healthcare Workers’ Employment Status and Mental Distress over SARS-CoV-2 in Bolivia

**DOI:** 10.1101/2023.08.07.23293747

**Authors:** Lea John, María Teresa Solís-Soto, Mira Mühlhäusser, Katja Radon

## Abstract

**Background:** Healthcare workers (HCW) have been particularly affected by the SARS-CoV-2 pandemic as it influenced employment conditions and unemployment/insecure employment. Their deterioration is associated with mental distress.

**Objective:** The aim of the study was to assess the trajectory of mental distress among HCW graduates during the COVID-19 pandemic in relation to their employment status.

**Methods:** We compared the change in mental distress over time among recent HCW graduates who were formally employed, to those who were unemployed/insecurely employed during the pandemic. In 2018 and 2022, we prospectively surveyed HCW who were in their final year of study in 2018 in Bolivia. Information was collected on socio-demographic characteristics, employment status, and mental distress. Mental distress was assessed using the 12-item General Health Questionnaire. Generalized Estimating Equations were implemented to examine changes in mental distress over time and the role of employment status in this development. Of the 663 HCW at baseline, 116 could be followed up.

**Findings:** Over the course of the pandemic, formal employment after graduation did not change the odds of mental distress (odds ratio (OR)=0.93 [95% confidence interval (CI) 0.13−6.83]). In contrast, unemployment/insecure employment statistically significantly increased the odds of mental distress (OR=2.10 [CI 1.05−4.24]) over time.

**Conclusions:** Especially in countries with limited social support for unemployed/insecurely employed citizens, interventions and policies to prevent mental distress among newly graduated HCW are important. This is particularly relevant in the face of crises such as the COVID-19 pandemic.

## Introduction

In January 2023, nearly three years after the Coronavirus disease 2019 (COVID-19) was declared as a global pandemic, the World Health Organization reported a cumulative number of nearly 661 million infections and 6.7 million deaths worldwide. In Bolivia, with the first confirmed case on March 12, 2020, 1.2 million cases of COVID-19 have been registered, and 22,324 people have died (1). COVID-19 is caused by the severe acute respiratory syndrome coronavirus 2 (SARS-CoV-2). The disease and the measures taken to contain its spread have had a major impact on daily life around the world. Particularly in low- and middle-income regions such as Latin America, the SARS-CoV-2 outbreak has caused major economic, social, political, and health crises (2).

The high prevalence of SARS-CoV-2 infections worldwide, its highly infectious nature, its severe course of the disease, and the measures taken to contain its spread have contributed to the development of mental distress in the population (3, 4). Mental distress is a major public health problem that reduces quality of life and increases mortality. According to a secondary analysis of the National UK Household Longitudinal Study (4) and a longitudinal study of US adults (3), mental distress increased significantly during the COVID-19 pandemic compared with pre-COVID-19 data (3, 4).

Among healthcare workers (HCW), the collapse of the healthcare system, long working hours, constant exposure to the virus, unavailability of personal protective equipment, as well as disrespect and violence towards HCW, played a fundamental role in the deterioration of mental health during the pandemic (2). It has been shown that younger HCW, who may still be in training or have recently graduated, have an increased likelihood of developing mental distress throughout the pandemic (2, 5, 6). No prospective studies, including pre-COVID-19 data from Latin America, were found to adress this issue. However, a review based on cross-sectional studies from 2022 concluded that the mental health of HCW worsened during the first year of the COVID-19 pandemic in Latin America (2).

In addition to the challenges described above, the COVID-19 pandemic has created massive employment insecurity both globally (7) and in Bolivia (8). Studies from Latin America have shown that employment insecurity and unemployment are significant risk factors for mental distress (9–11), especially during the SARS-CoV-2 pandemic (7, 8). Likewise, formal employment as a new healthcare graduate in Mexico during the COVID-19 pandemic was found to be a protective factor against mental distress. Prior to the pandemic, students’ risk of mental distress was shown to decrease as they entered their professional career (12). However, no studies were found that examined this development during the COVID-19 pandemic.

Forms of insecure employment include informal employment and underemployment (13). Informal employment is characterized by low skill requirements and economic barriers to entry, labor-intensive production in small family businesses, and an unregulated and competitive market (10, 13). Underemployment occurs when individuals are employed but not according to their skills, are underpaid, or work fewer hours than desired (14). Unemployment, on the other hand, occurs when individuals do not have a job that pays a regular salary (15). In Bolivia, unemployment and underemployment rates have increased during the COVID-19 pandemic and have begun to decline again since the end of 2021: unemployment rates were 4% in 2019, 7% in 2021, and 4% in 2022, and underemployment rates were 4% in 2019, 10% in January 2021 and 7% in December 2021 (16). In Bolivia, weak social policies have resulted in a lack of social benefits for the unemployed and underemployed, leading to high informal employment rates of 85% in 2019 and estimated 90% in 2021 (10, 17, 18). Unemployment and insecure employment particulary affect young adults who have recently graduated (7). In the healthcare sector, financial support from international organizations has helped improve access and treatment for the Bolivian population in recent decades. However, the Bolivian health sector remains underfunded, resulting in a shortage of supplies, facilities and qualified HCW (19). Young HCW, in particular, face an employment crisis associated with job insecurity, unemployment, informal employment, and underemployment characterized by low-income and low-skill work (18, 20).

In summary, the working conditions in the healthcare sector worsened worldwide during the COVID-19 pandemic (2). In addition to the difficult employment situation of HCW in Bolivia before the pandemic, insecure employment and unemployment increased significantly (10, 13, 16, 17), particularly among young adults (7). Both of these factors may increase mental distress (2, 9–11). The question arises as to how these conditions affect the − under normal circumstances − positive development of students’ mental health as they enter professional careers. Therefore, we followed up recent HCW graduates in Sucre, Bolivia to examine how the prevalence of mental distress changed since the onset of the SARS-CoV-2 pandemic and whether employment status after graduation (i.e., formal employment vs. unemployment/insecure employment) played a role in this development.

## Materials and Methods

### Study Design

A prospective cohort study was conducted beginning in 2018. Nursing and medical students at the Universidad San Francisco Xavier de Chuquisaca (USFX) Sucre, Bolivia were recruited. Participants were in their last year of academic training before the internship (third year in the career of nursing and fifth year in the career of medicine), over 18 years of age, and Spanish-speaking (21). A total of 663 out of 809 invited students (82%) participated in the baseline study. Participants were followed up in 2022.

For the baseline study, study coordinators visited students’ classes and explained the objectives and procedures of the study. Participants were given a tablet to complete the digital questionnaire on site. For the follow-up study, emails, and WhatsApp messages with a link to the questionnaire were distributed to participants who had agreed to be contacted again at baseline (n = 526, 79%). A reminder was sent after one, three, and four weeks after the initial mailing to ensure a high response rate. As an incentive, participants who completed the questionnaire received five Farmacorp (Farmacy) vouchers worth 200 Bolivianos (approx. 29 USD) each, which were distributed through a lottery. To further promote the follow-up study, it was announced on the USFX University television and radio as well as on Bolivia’s public radio station ’La Bruja’. In the end, 116 (22%) participants completed the follow-up questionnaire (Figure A1).

The survey was pseudo-anonymous, and participants could withdraw from the study at any time without giving a reason. All participants gave written informed consent to participate in the baseline and follow-up study. The study was approved by the Ethics Committee of the Faculty of Medicine at the Universidad Mayor de San Simon (no project number assigned; November 8, 2021) and by the Ethics Committee of the Faculty of Medicine at the Ludwig-Maximilians Universität in Munich (project number 22-0451; June 11, 2022).

### Questionnaire Instruments and Variable Definition

The baseline questionnaire included 44 questions to assess the participants’ socio-demographic characteristics, mental distress, lifestyle, employment status, working conditions, academic training, coping strategies, and social support. The questions were provided in Spanish and were administered as an online questionnaire using SurveyMonkey (Momentive Europe UC, Dublin, Ireland) at baseline and follow-up.

Employment status and socio-demographic characteristics were assessed using parts of the Quality of Life and Employment, Labor, and Health Conditions Questionnaire of the First National Survey of Workers in Chile (ENETS) (22) and parts of the Job Prospects and Employability in Nursing and Psychology Questionnaire (23) developed in Mexico.

Employment status was assessed as exposure. It had to be defined differently at baseline and follow-up, as participants had not yet completed the HCW training at baseline. At baseline, employment was defined as a positive response to the question ’Are you currently working?’. At follow-up, participants were considered employed as HCW if they answered ’yes’ to the two questions ’Are you currently working?’ and ’Is your work related to your profession?’. If the answer to wither question was ’no’, the participant was considered ’unemployed or employed insecurely’. In addition, those who were formally employed at follow-up were asked if they worked with COVID-19 patients. Those who were unemployed or insecurely employed at follow-up were asked why they were not formally employed (options: ’I stopped looking for a job because I couldn’t find one’, ’I am currently studying or in training’ or ’other reasons’).

Socio-demographic characteristics included sex, age, marital status, economic situation, and whether participants had children. Because sex and econimic situation are likely to be associated with both exposure (employment status) and outcome (mental distress), they were considered as potential confounders (4, 7). Age was not included as a potential confounder because of its limited range (97% 19−30 years).

Mental distress was assessed using the validated Spanish version of the 12-item General Health Questionnaire (GHQ-12) (24). The GHQ-12 was originally developed in English by Goldberg and colleagues. It is a well-established self-report measure of mental distress designed to screen for psychological morbidity and psychiatric disorders in community and non-psychiatric settings. On a 4-point scale from ’much more than usual’, ’rather more than usual’, ’no more than usual’ to ’not at all’, participants indicated how often they had experienced various psychological symptoms in the past four weeks. The outcome was a binary threshold score on the GHQ-12. The threshold measure was scored using the GHQ scoring method with 0-0-1-1 for positive items and 1-1-0-0 for negative items, and summed (range 0–12) (25). As suggested by a Chilean validation study (26), participants with a GHQ-12 score > 4 (threshold score 4/5) were defined as having a clinically relevant level of mental distress (hereafter referred to as ’mental distress’).

### Statistical Analysis

Data analysis was performed using SPSS version 28.0 (IBM, Armonk, NY, USA). To assess selective losses in the follow-up study, we compared subjects who participated in both studies with those who dropped out. Differences between these two groups were examined using chi-square tests for binary variables and paired t-tests for continuous variables. To examine systematic differences between being employed as an HCW and being unemployed or insecurely employed as an HCW, we compared these groups at follow-up. Nominal and ordinal variables were described as absolute and relative frequencies. Metric variables were described as mean and standard deviation (SD). We used chi-square tests to assess the association between socio-demographic characteristics and the employment status with mental distress in bivariate analysis.

Missing data on outcome, exposure, and potential risk factors were multiply imputed (n = 20) for participants who answered at least one question in the baseline and follow-up questionnaires. Assuming that these data were missing at random, the Fully Conditional Specification (FSC) method was used with the Predictive Mean Matching (PMM) scaling model, as implemented in SPSS software (27).

To analyze associations between exposure and outcome, Generalized Estimating Equations (GEE) were used to estimate odds ratios (OR) and corresponding 95% confidence intervals (95% CI) using the logit link function. Participants were used as the clustering factor. Time (T0 = baseline, T1 = follow-up) was considered as an inner subject effect. First, unadjusted results were estimated for time, exposure (employment status), and all potential risk factors. In the next step, the model was adjusted for the potential confounders sex and economic situation. Associations of change in mental distress with employment over time were analyzed by including an interaction term between time and employment status. An interaction was suspected because the employment status changed between T0 and T1 due to the participants’ graduation in the meantime and the influence of the COVID-19 pandemic (7, 8, 28).

Several sensitivity analyses were performed. First, we used a threshold score of 5/6 for the binary threshold score of the GHQ-12. Second, the continuous mean GHQ-12 score was used as the outcome. For this, the GHQ-12 items were scored according to the Likert scoring method (0-1-2-3 for positive items and inverted for negative items), resulting in a score of 0–36 (29). In the GEE, the identity link function was used to analyze the continuous outcome. Third, we changed the definition of the exposure variable (employment status) at baseline. Employment was defined the same as at follow-up as an affirmative response to the question, ’Is your work related to your profession?’. Fourth, those who were still in academic training at follow-up were excluded from the analysis. Finally, fully imputed data were analyzed, taking into account all those who participated at baseline.

## Results

At baseline, the mean age of the 663 participants was 23.9 (SD 2.9) years. More women (68%) than men (32%) and more medical students (80%) than nursing students (20%) participated in the study.

There was no statistically significant difference between participants who remained in the study and those who dropped out (p > 0.05 for all variables). However, while 26% of those who remained in the study were employed at baseline, only 20% of those lost to follow-up did so. The prevalence of mental distress at baseline was similar in the two groups (64% of retained versus 65% of lost to follow-up, Table 1).

**Table 1.** Description of the study population stratified for non-responders and responders (N = 663).

| Characteristics |  | No.<br>missing<br>(%) | Baseline participants |  |  |
| --- | --- | --- | --- | --- | --- |
|  |  |  | N = 663 |  |  |
|  |  |  | Non-Responders | Responders | P |
|  |  |  | n = 547 | n = 116 |  |
|  |  |  | n (%)<sup>0</sup> | n (%)<sup>0</sup> |  |
| Sex | Female | 0 | 364 (66.5) | 88 (75.9) | 0.062 |
| Marital status in 2018 | Single<sup>+</sup> | 0 | 444 (81.2) | 91 (78.4) | 0.518 |
| Career | Medicine<sup>#</sup> | 0 | 447 (81.7) | 86 (74.1) | 0.071 |
| Economic situation in 2018 | Good |  | 119 (22.3) | 29 (25.4) | 0.665 |
|  | Neither good nor bad | 15 (2.3) | 343 (64.2) | 68 (59.6) |  |
|  | Bad |  | 72 (13.5) | 17 (14.9) |  |
| Children in 2018 | Yes | 0 | 95 (17.4) | 18 (15.5) | 0.685 |
| Employment in 2018* | Yes | 2 (0.3) | 110 (20.2) | 30 (25.9) | 0.210 |
| Mental distress in 2018 | Yes | 5 (0.8) | 347 (64.0) | 75 (64.7) | 0.916 |
|  |  |  | Mean (SD) | Mean (SD) |  |
| Age (years) in 2018 |  | 0 | 24.0 (0.1) | 23.5 (0.3) | 0.085 |

Non-Responders: those who did not participate in the follow-up study; Responders: those who remained in the study throughout the two time periods; Mental distress: 12-item General Health Questionnaire threshold above 4 indicates a clinically relevant level of mental distress (range 0–12); <sup>0</sup>% of non-missing data; <sup>+</sup>Otherwise 'married or having a partner'; <sup>#</sup>Otherwise 'nursing'; \*Participants were still in academic training (medicine or nursing); SD: standard deviation; Chi-square test for binary variables, paired t-test for continuous variable); Non-imputed data.

At follow-up, 49% of the population worked as HCW in formal employment. Of the unemployed or insecurely employed HCW, 32% were currently studying or undergoing training. Overall, formally employed HCW at follow-up were more likely to be in an economically disadvantaged situation at baseline compared to those HCW being unemployed or insecurely employed at follow-up (22% vs. 17%, Table 2). It was slightly more frequent among formally employed HCW at follow-up to be single than among unemployed or insecurely employed HCW (78% vs 76%). Forty-one (80%) formally employed HCW reported having worked with COVID-19 patients.

**Table 2.**
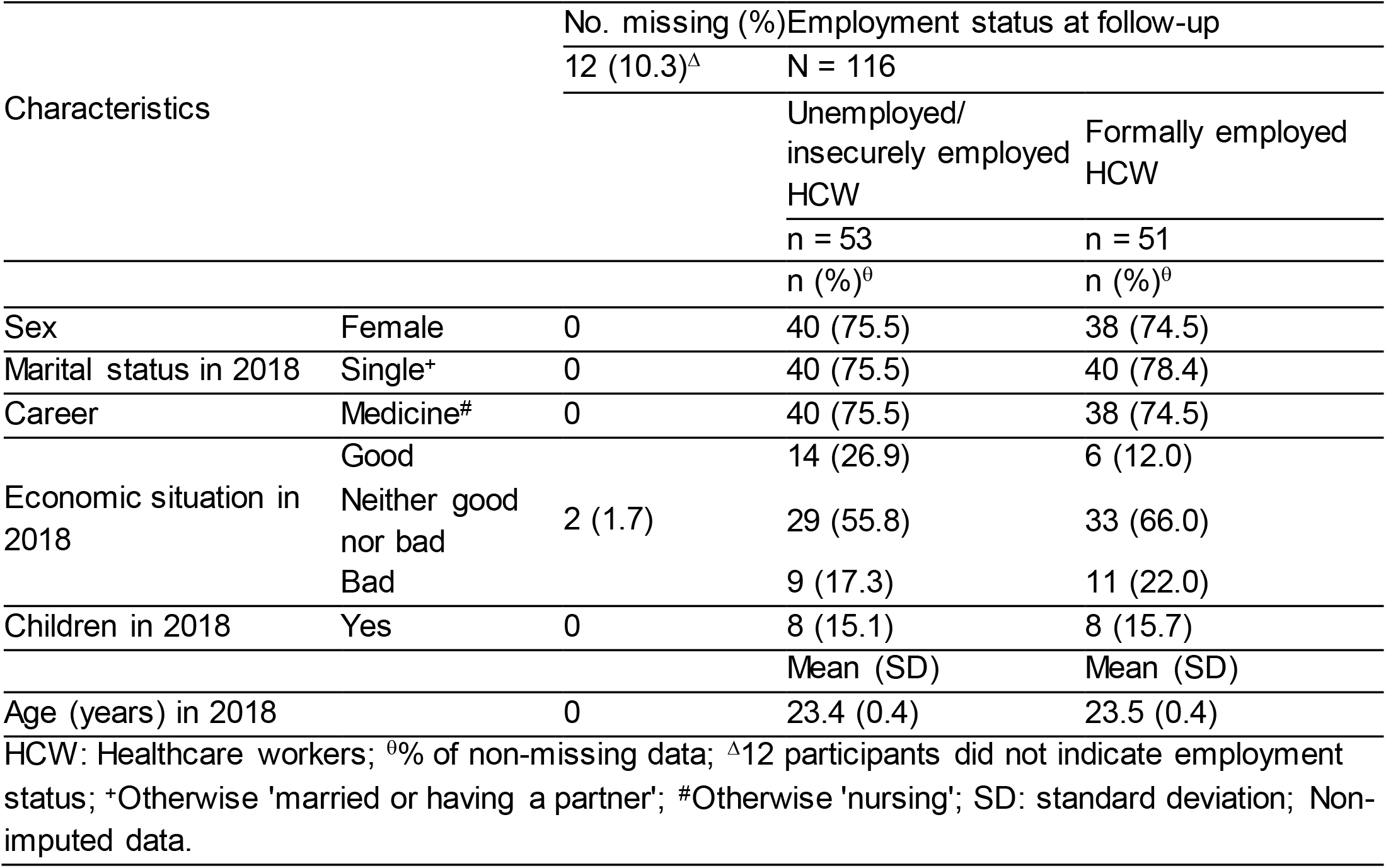
Description of the study population stratified for participants who were unemployed/insecurely employed as HCW or were formally employed as HCW at follow-up (N = 116).

Restricting the study population to those who answered the GHQ-12 questions at baseline and follow-up (n = 103), the prevalence of mental distress increased from 65% at baseline to 69% at follow-up, which was not statistically significant.

At baseline, being female and having a less favorable economic situation were statistically significantly associated with mental distress. These associations were not found at follow-up. Among HCW who were employed at baseline the prevalence of mental distress decreased over time (70% vs. 64%). In contrast, among HCW who were employed at follow-up, the prevalence of mental distress did not change between baseline (59%) and follow-up (59%, Table 3).

**Table 3.**
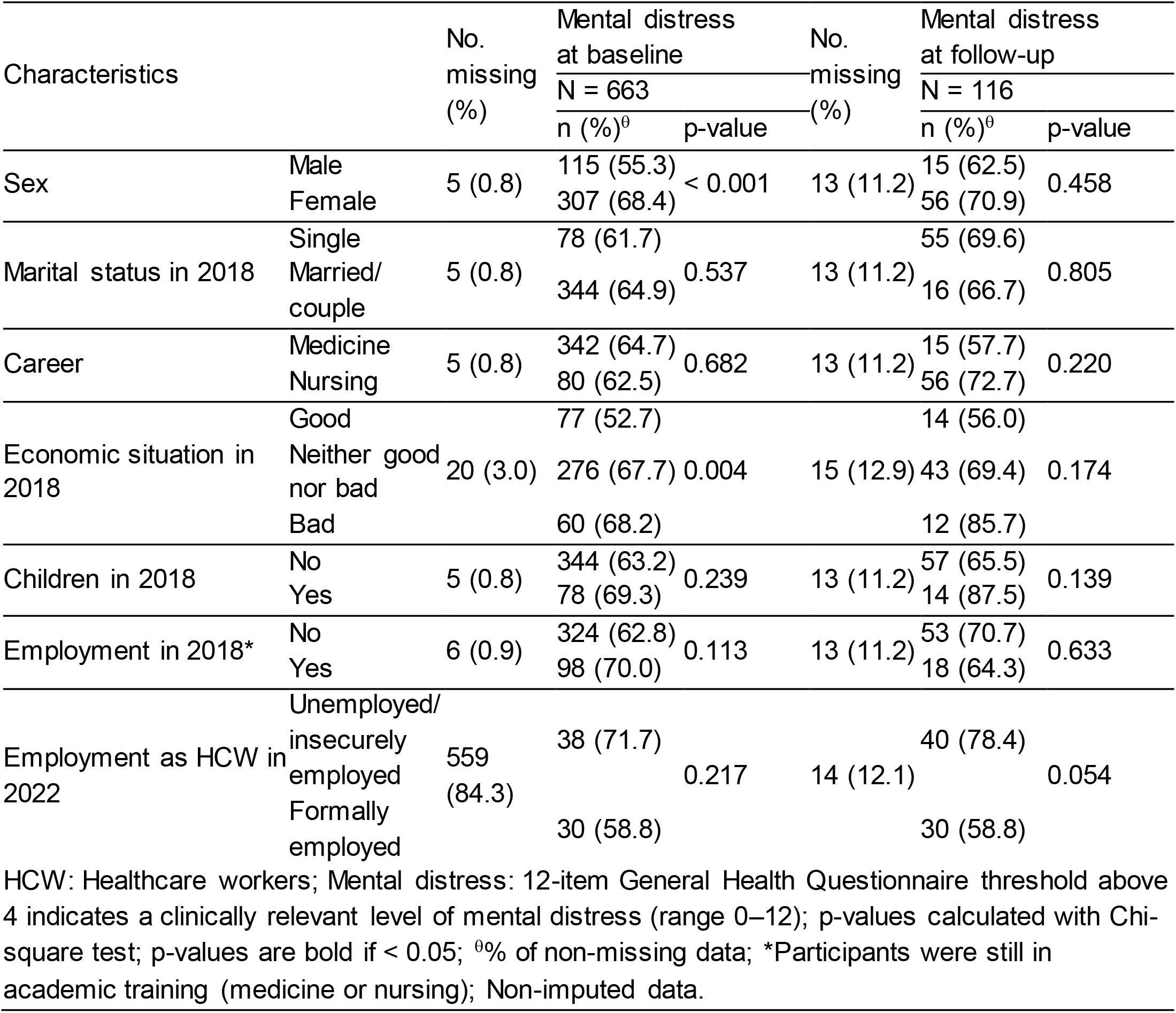
Prevalence of mental distress among participants at baseline (N = 663) and follow-up (N = 116) by potential risk factors.

| Characteristics |  | No.<br>missing<br>(%) | Mental distress<br>at baseline |  | No.<br>missing<br>(%) | Mental distress<br>at follow-up |  |
| --- | --- | --- | --- | --- | --- | --- | --- |
|  |  |  | N = 663 |  |  | N = 116 |  |
|  |  |  | n (%)<sup>0</sup> | p-value |  | n (%)<sup>0</sup> | p-value |
| Sex | Male | 5 (0.8) | 115 (55.3) | < 0.001 | 13 (11.2) | 15 (62.5) | 0.458 |
|  | Female |  | 307 (68.4) |  |  | 56 (70.9) |  |
| Marital status in 2018 | Single | 5 (0.8) | 78 (61.7) | 0.537 | 13 (11.2) | 55 (69.6) | 0.805 |
|  | Married/<br>couple |  | 344 (64.9) |  |  | 16 (66.7) |  |
| Career | Medicine | 5 (0.8) | 342 (64.7) | 0.682 | 13 (11.2) | 15 (57.7) | 0.220 |
|  | Nursing |  | 80 (62.5) |  |  | 56 (72.7) |  |
| Economic situation in<br>2018 | Good | 20 (3.0) | 77 (52.7) | 0.004 | 15 (12.9) | 14 (56.0) | 0.174 |
|  | Neither good<br>nor bad |  | 276 (67.7) |  |  | 43 (69.4) |  |
|  | Bad |  | 60 (68.2) |  |  | 12 (85.7) |  |
| Children in 2018 | No | 5 (0.8) | 344 (63.2) | 0.239 | 13 (11.2) | 57 (65.5) | 0.139 |
|  | Yes |  | 78 (69.3) |  |  | 14 (87.5) |  |
| Employment in 2018* | No | 6 (0.9) | 324 (62.8) | 0.113 | 13 (11.2) | 53 (70.7) | 0.633 |
|  | Yes |  | 98 (70.0) |  |  | 18 (64.3) |  |
| Employment as HCW in<br>2022 | Unemployed/<br>insecurely<br>employed | 559<br>(84.3) | 38 (71.7) | 0.217 | 14 (12.1) | 40 (78.4) | 0.054 |
|  | Formally<br>employed |  | 30 (58.8) |  |  | 30 (58.8) |  |

HCW: Healthcare workers; Mental distress: 12-item General Health Questionnaire threshold above 4 indicates a clinically relevant level of mental distress (range 0–12); p-values calculated with Chi-square test; p-values are bold if < 0.05; <sup>0</sup>% of non-missing data; \*Participants were still in academic training (medicine or nursing); Non-imputed data.

The GEE models included all available data on participants (N = 663 for baseline and N = 116 for follow-up). Consistent with the unadjusted results, employment at baseline was not statistically significantly associated with mental distress in the adjusted model (OR=1.29 [95% CI 0.86−1.93]). At follow-up formal employment as an HCW was not statistically significantly associated with mental distress (OR=0.93 [95% CI 0.13−6.83]). After adjustment, the odds of mental distress increased statistically significantly from baseline to follow-up for unemployed or insecurely employed HCW in the adjusted model (OR=2.10 [95% CI 1.05−4.24], Table 4).

**Table 4.** Results of Generalized Estimating Equations models for mental distress with threshold score 4/5 between baseline and follow-up.

| Characteristics |  | Mental distress |  |
| --- | --- | --- | --- |
|  |  | N = 663<sup>0</sup> |  |
|  |  | Crude OR<br>(95% CI) | Adjusted OR<br>(95% CI) |
| Sex | Male | 1 | 1 |
|  | Female | 1.71 (1.29–2.37) | 1.68 (1.21–2.34) |
| Economic situation | Good | 1 | 1 |
|  | Neither good nor bad | 1.65 (1.16–2.35) | 1.53 (1.07–2.20) |
|  | Bad | 2.02 (1.19–3.43) | 1.92 (1.12–3.29) |
| Employment | No | 1 | 1 |
|  | Yes | 1.12 (0.78–1.60) | 1.29 (0.86–1.93) |
| Study phase | Baseline: 2018 | 1 | 1 |
|  | Follow-up: 2022 | 1.31 (0.86–2.00) | 2.10 (1.05–4.24) |
| Interaction | Not employed at baseline* | 1 | 1 |
|  | Formally employed as<br>HCW at follow-up | 0.93 (0.16–5.28) | 0.93 (0.13–6.83) |

HCW: Healthcare workers; Mental distress: 12-item General Health Questionnaire threshold above 4 indicates a clinically relevant level of mental distress (range 0–12); Estimates are bold if CI excludes 1; <sup>0</sup>Follow-up data available for N = 116; Adjusted for sex and economic situation; \*Participants were still in academic training (medicine or nursing); Imputed data.

The results of all sensitivity analyses confirmed our findings. In all GEE models, mental distress increased among unemployed and insecurely employed HCW during the COVID-19 pandemic. However, the results were no longer statistically significant. In all GEE models, mental distress did not change among formally employed HCW during the COVID-19 pandemic (Tables A1-A5).

## Discussion

This study is the first to track employment and mental distress among HCW who were in their final year of study in 2018, just before the outbreak of the COVID-19 pandemic in Bolivia. Contrary to expectations from pre-COVID-19 studies, the prevalence of mental distress among medical and nursing students (T0) and graduates (T1) did not decrease during the SARS-CoV-2 pandemic, nor did it increase overall. However, we found a statistically significant increase in mental distress among unemployed and insecurely employed HCW three years into the pandemic. In contrast, among those formally employed as medical or nursing graduate, there was no statistically significant change in mental distress during the pandemic.

### Consistency with other Studies

Comparing our findings with other studies of mental distress in Bolivia, we found that young HCW had a higher prevalence of mental distress than Bolivian teachers (43%) and a lower prevalence than Bolivian miners (81%) (30, 31). Given the extreme working conditions, such as very high and very low temperatures, noise, and shift work, the higher prevalence of mental distress among minders is not surprising (31). The comparatively lower prevalence of mental distress among teachers may be due to the use of a higher threshold score of 5/6 for the GHQ-12, which results in a lower estimate of mental distress (30). However, using a threshold score of 5/6 in our sensitivity analyses did not change the association between the employment status and mental distress (Table A1). The difference in prevalence could also be attributable to the impact of the COVID-19 pandemic on HCW. As pointed out by Ferreira, L.C. et al. (32), graduates faced unprecedented challenges related to the pandemic, such as changes in internships, loss of space for practice scenarios, as well as cancellation of graduation rites and ceremonies. Participants entered a highly demanding workplace and, simultaneously, a work environment with immense employment insecurity (32). These conditions can lead to a high prevalence of mental distress (7, 8, 28). Comparing our results with the COVID-19 Healthcare Workers Study (HEROES), a prospective cohort study evaluating the impact of the COVID-19 pandemic on the mental health of HCW in 26 LMIC and HIC, including Bolivia (33), the mean GHQ-12 scores were higher in our study (34). This heterogeneity in results may stem from the older age of participants in the HEROES study compared with our study population, as young HCW are more likely to develop mental distress (2, 5, 6).

Prior to the pandemic, students were shown to be at increased risk for mental distress (35). Consequently, under normal circumstances, mental distress decreases again with the onset of the professional career (12). However, as a result of the COVID-19 pandemic, our results showed that mental distress among young HCW did not decrease with entry into formal employment but has remained at a consistently high level. Our findings are supported by several studies that found an increase in mental distress among HCW over the course of the COVID-19 pandemic (2, 36, 37). Other studies contradict these findings (38, 39). Possible reasons for these inconsistent findings include differences in the operationalization of mental distress, cultural factors, and differences in the study population with respect to age, sex and occupation.

Among unemployed and insecurely employed HCW the prevalence of mental distress increased after graduation during the COVID-19 pandemic. This finding is consistent with other studies (6, 7, 40): In a secondary analysis of a longitudinal population-based study it was found that unemployment and insecure employment negatively affected mental health in the general population of the United States during the SARS-CoV-2 pandemic (7). Medina Fernández, I.A. et al. conducted a cross-sectional study of the mental health of Mexican graduate students in the context of the COVID-19 pandemic. Consistent with our results, they found that those who work as HCW were less likely to experience mental distress than those who did not work as HCW during the COVID-19 pandemic (6). Likewise, another cross-sectional study investigating the mental health of recentuniversity graduates in Brazil found that graduates were less affected by mental distress when they were formally employed (40).

However, a study frm the United Kingdom found different results: Pierce, M. et al. examined the changes in adult mental health in the general population before and one month into the COVID-19 pandemic. They found an increase in the mean GHQ-12 score in the employed population and no change in the mean GHQ-12 score in the unemployed population (4). However, it must be said that the results may not be comparable to ours for the following reasons: First, the general population was examined, not recently graduated HCW. Second, insecure employment has fundamentally different consequences in the UK (and other HICs) than in Bolivia (and other LMICs). Without a social welfare system to support the unemployed and insecurely employed, having a secure and well-paying job is essential for survival in Bolivia (10, 13). These circumstances influence the impact of the employment status on mental distress (9–11). In addition, the initial increase in mental distress one month into the pandemic found by Pierce, M. et al. may represent a spike in emotional response that stabilizes or declines as people adjust (4). The present study reported the long-term effects of the pandemic on participants’ mental health status three years into the pandemic. At this advanced stage of the pandemic, HCW may have gained resilience (38, 39).

### Limitations and Strengths

Although we had a large group of participants at baseline with a high response rate (82%), the response rate at follow-up was comparatively low (22%). Therefore, our results cannot be considered representative of the study population. Therefore, selection bias may be introduced. However, the non-responder analysis showed that participants who were lost to follow-up were not statistically significantly different from responders who remained in the study at both time points. Nevertheless, low statistical power at follow-up may have resulted in failure to detect changes in important factors between baseline and follow-up.

We recognize that our exposure definition differs between baseline and follow-up because employment status varies depending on whether participants were still in training (T0) or were trained HCW (T1). This needs to be taken into account when using ’not employed at baseline’ as reference group. There may also be other aspects that changed between T0 and T1, such as participants moving out of Sucre, Bolivia after graduation. However, sensitivity analysis showed that the association between employment status and mental distress did not change when employment at baseline was defined identically to employment at follow-up as employment as HCW (Table A2). Excluding participants at follow-up who stated to still being in training also did not change the association (Table A3).

Misclassification of exposure and outcome may have occurred because they were assessed only by questionnaire. The GHQ-12 is a validated screening tool that correlates strongly with the presence of mental illness and with future clinical diagnosis of a psychiatric disorder. However, the GHQ-12 is not equivalent to a clinical diagnosis (41). This study relied on self-report, which is valid primarily because the effect of employment status on mental distress depends largely on personal assessment of the situation. This leads us to the conclusion that the possibility of measurement error is low (42). Bias in the negatively worded items of the GHQ-12 may have occurred due to inattentive reading by responders, i.e., the respondent did not notice that the response format had changed (positive and negative items) (43).

This study cannot conclusively identify the reasons why economically disadvantaged participants are more likely to be formally employed than participants in an economically advantaged situation. In addition to policy reasons, explanations could be that participants with greater economic needs applied at an early stage for formal employment and did not, e.g., complete further training. Early entry into employment allowed them to develop soft- and work-related skills, which made them more attractive to the formal labor market.

The present study did not distinguish between the different forms of underemployment and unemployment. Underemployment can be income-, skill-or hours-based (14). Whereas unemployment can be frictional, structural, or cyclical (44). Likewise, we combined unemployed and insecurely employed HCW into one group because there is evidence that insecurely employed people mirror unemployed people in terms of their mental health (7). We also did not distinguish between frontline HCW, which are HCW providing direct care for COVID-19 patients, and normal HCW, although there is indication that the prevalence may be higher among frontline HCW (45). Consequently, a dose-response relationship could not be assessed.

Due to the prospective design of the study the risk of recall bias was limited because the temporal sequence of exposure and outcome could be evaluated. Nevertheless, the possibility of reverse causation cannot be excluded. For example, childhood mental health problems could not be included as a potential confounder because such information was not available. Potential confounding by sex and economic situation was taken into account, although we did not consider aspects such as the impact of measures such as home-schooling, other occupations, such as caring for relatives, whether participants had been diagnosed with COVID-19 or other serious and/or chronic diseases, and whether participants had been vacinated against the SARS-CoV-2 virus (7).

## Conclusion

Our study describes an increase in mental distress among unemployed and insecurely employed HCW after graduation during the SARS-CoV-2 pandemic, while no change was observed among those who were formally employed. Particularly in countries with a labor market dominated by informal employment, underemployment, and unemployment with limited social support for citizens, such as Bolivia, it may be useful to consider these findings. Interventions and a change in policy strategies are needed to prevent mental distress among young HCW, especially in the face of crises such as the COVID-19 pandemic.

## Data Availability

All data produced in the present study are available upon reasonable request to the authors.

---

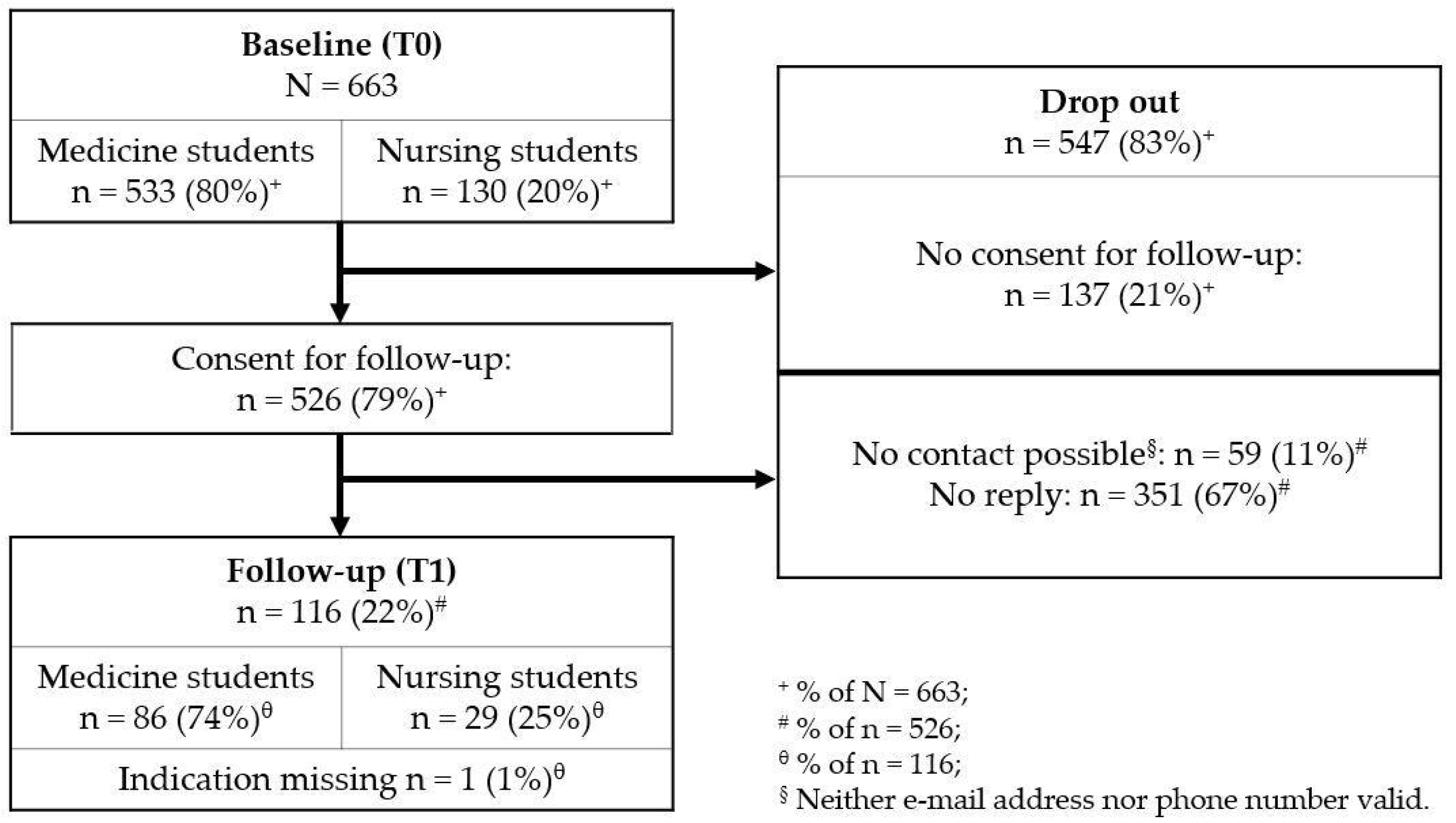

