## Supplementary Tables A1-A5 for "Young Healthcare Workers’ Employment Status and Mental Distress over SARS-CoV-2 in Bolivia"

Table A1. Sensitivity analysis: Results of Generalized Estimating Equations models for mental distress with threshold score 5/6 between baseline and follow-up.

| Characteristics | | Mental distress N = 663$^{\theta}$ Crude OR (95% CI) | Mental distress N = 663$^{\theta}$ Adjusted OR (95% CI) |
|---|---|---|---|
| Sex | Male | 1 | 1 |
| | Female | 1.13 (1.04–1.22) | 1.62 (1.17–2.24) |
| Economic situation | Good | 1 | 1 |
| | Neither good nor bad | 1.05 (0.96–1.15) | 1.15 (0.80–1.65) |
| | Bad | 1.17 (1.04–1.32) | 1.85 (1.10–3.09) |
| Employment | No | 1 | 1 |
| | Yes | 1.02 (0.72–1.44) | 1.10 (0.75–1.62) |
| Study phase | Baseline: 2018 | 1 | 1 |
| | Follow-up: 2022 | 1.31 (0.88–1.96) | 1.70 (0.93–3.14) |
| Interaction | Not employed at baseline* | 1 | 1 |
| | Formally employed as HCW at follow-up | 1.05 (0.11–9.78) | 1.03 (0.15–6.86) |

HCW: Healthcare workers; Mental distress: 12-item General Health Questionnaire threshold above 5 indicates a clinically relevant level of mental distress (range 0–12); Estimates are bold if CI excludes 1; $^{\theta}$Follow-up data available for N = 116; Adjusted for sex and economic situation; *Participants were still in academic training (medicine or nursing); Imputed data.

Table A2. Sensitivity analysis: Results of Generalized Estimating Equations models for mental distress with threshold score 4/5 between baseline and follow-up with identical exposure definition at both time points.

| Characteristics | | Mental distress N = 663[θ] Crude OR (95% CI) | Adjusted OR (95% CI) |
|---|---|---|---|
| Sex | Male | 1 | 1 |
| | Female | 1.36 (0.56–3.31) | 1.34 (0.62–2.89) |
| Economic situation | Good | 1 | 1 |
| | Neither good nor bad | 1.44 (0.90–2.10) | 1.39 (0.96–2.01) |
| | Bad | 1.83 (0.91–3.69) | 1.79 (0.92–3.42) |
| Employment | No | 1 | 1 |
| | Yes | 0.89 (0.32–2.47) | 1.01 (0.46–2.20) |
| Study phase | Baseline: 2018 | 1 | 1 |
| | Follow-up: 2022 | 1.75 (0.73–4.17) | 2.06 (0.66–6.46) |
| Interaction | Unemployed/insecurely employed at baseline* | 1 | 1 |
| | Formally employed as HCW at follow-up | 1.65 (0.20–13.52) | 1.79 (0.14–22.42) |

HCW: Healthcare workers; Mental distress: 12-item General Health Questionnaire threshold above 4 indicates a clinically relevant level of mental distress (range 0–12); Estimates are bold if CI excludes 1; [θ]Follow-up data available for N = 116; Adjusted for sex and economic situation; *Employment was defined the same as at follow-up as an affirmative response to the question 'Is your work related to your profession?'; Imputed data.

Table A3. Sensitivity analysis: Results of Generalized Estimating Equations models for mental distress with threshold score 4/5 between baseline and follow-up, excluding those studying or being in training at follow-up.

| Characteristics | | Mental distress N = 663[θ] Crude OR (95% CI) | Adjusted OR (95% CI) |
|---|---|---|---|
| Sex | Male | 1 | 1 |
| | Female | 1.71 (1.23–2.38) | 1.67 (1.20–2.33) |
| Economic situation | Good | 1 | 1 |
| | Neither good nor bad | 1.71 (1.19–2.45) | 1.58 (1.09–2.28) |
| | Bad | 2.07 (1.21–3.54) | 1.94 (1.12–3.35) |
| Employment | No | 1 | 1 |
| | Yes | 1.09 (0.76–1.57) | 1.29 (0.86–1.94) |
| Study phase | Baseline: 2018 | 1 | 1 |
| | Follow-up: 2022 | 1.38 (0.87–2.21) | 3.37 (1.20–9.46) |
| Interaction | Not employed at baseline* | 1 | 1 |
| | Formally employed as HCW at follow-up | 0.91 (0.12–7.03) | 0.91 (0.09–8.76) |

HCW: Healthcare workers; Mental distress: 12-item General Health Questionnaire threshold above 4 indicates a clinically relevant level of mental distress (range 0–12); Estimates are bold if CI excludes 1; [θ]Follow-up data available for N = 100 (excluding n = 16, who were studying or being in training at follow-up); Adjusted for sex and economic situation; *Participants were still in academic training (medicine or nursing); Imputed data.

Table A4. Sensitivity analysis: Results of Generalized Estimating Equations models for mental distress with mean GHQ-12 score between baseline and follow-up.

| Characteristics | | Mental distress N = 663$^{\theta}$ Crude OR (95% CI) | Mental distress N = 663$^{\theta}$ Adjusted OR (95% CI) |
|---|---|---|---|
| Sex | Male | 0 | 0 |
| | Female | 1.78 (0.79–2.76) | 1.76 (0.79–2.72) |
| Economic situation | Good | 0 | 0 |
| | Neither good nor bad | 0.99 (-0.13–2.12) | 0.82 (-0.30–1.94) |
| | Bad | 2.63 (1.09–4.17) | 2.56 (1.02–4.10) |
| Employment | No | 0 | 0 |
| | Yes | -0.17 (-1.19–0.86) | 0.09 (-1.02–1.20) |
| Study phase | Baseline: 2018 | 0 | 0 |
| | Follow-up: 2022 | 1.07 (-0.75–2.21) | 2.04 (0.48–3.61) |
| Interaction | Not employed at baseline* | 0 | 0 |
| | Formally employed as HCW at follow-up | 0.11 (-2.56–2.80) | -0.04 (-3.18–3.10) |

HCW: Healthcare workers; Mental distress: Higher scores of the mean 12-item General Health Questionnaire score (range 0–36) representing a higher level of mental distress; Estimates are bold if CI excludes 0; $^{\theta}$Follow-up data available for N = 116; Adjusted for sex and economic situation; *Participants were still in academic training (medicine or nursing); Imputed data.

Table A5. Sensitivity analysis: Results of Generalized Estimating Equations models for mental distress with threshold score 4/5 between baseline and follow-up with full data imputation.

| Characteristics | | Mental distress N = 663$^{\theta}$ Crude OR (95% CI) | Adjusted OR (95% CI) |
|---|---|---|---|
| Sex | Male | 1 | 1 |
| | Female | 1.43 (0.58–3.56) | 1.43 (0.61–3.33) |
| Economic situation | Good | 1 | 1 |
| | Neither good nor bad | 1.34 (0.82–2.19) | 1.27 (0.80–2.03) |
| | Bad | 1.54 (0.67–3.58) | 1.47 (0.67–3.25) |
| Employment | No | 1 | 1 |
| | Yes | 1.08 (0.66–1.78) | 1.33 (0.88–2.02) |
| Study phase | Baseline: 2018 | 1 | 1 |
| | Follow-up: 2022 | 1.81 (0.73–4.50) | 2.22 (0.77–6.43) |
| Interaction | Not employed at baseline* | 1 | 1 |
| | Formally employed as HCW at follow-up | 1.70 (0.24–11.82) | 1.78 (0.21–15.18) |

HCW: Healthcare workers; Mental distress: 12-item General Health Questionnaire threshold above 4 indicates a clinically relevant level of mental distress (range 0–12); $^{\theta}$Complete data available for baseline and follow-up; Adjusted for sex and economic situation; *Participants were still in academic training (medicine or nursing); Imputed data.
